# Validation of 3D-DXA–Derived Proximal Femur Measurements Against QCT Across International Clinical Cohorts

**DOI:** 10.64898/2026.04.22.26351450

**Authors:** Marta I. Bracco, Dennis M. Black, Teruki Sone, Luis del Rio, Silvana de Gregorio, Jorge Malouf, Ludovic Humbert

## Abstract

Three-dimensional dual-energy X-ray absorptiometry (3D-DXA) reconstructs proximal femur models from standard scans to estimate cortical and trabecular bone parameters. The aim of this study was to evaluate 3D-DXA against quantitative computed tomography (QCT) across independent international cohorts. The study included 537 subjects from four cohorts: an adult population from Spain, a postmenopausal female population from the United States, an osteoarthrosis population and a young population, both from Japan. Subjects underwent both 3D-DXA and QCT imaging. Accuracy was assessed using linear regression and Bland-Altman analysis to evaluate systematic and random errors. 3D-DXA parameters strongly correlated with QCT across all datasets, with correlation coefficients between 0.82 and 0.97. Random errors were consistent across cohorts and ranged between 16.55 and 19.91 mg/cm^3^ for integral volumetric bone mineral density (vBMD), between 13.52 and 18.47 mg/cm^3^ for trabecular vBMD, and between 9.13 and 11.37 mg/cm^2^ for cortical surface bone mineral density (sBMD). Systematic errors ranged between -14.84 and 4.50 mg/cm^3^ for integral vBMD, between -8.31 and 14.41 mg/cm^3^ for trabecular vBMD, and between -5.58 and 3.21 mg/cm^2^ for cortical sBMD. The variations in systematic errors were likely attributable to differences in QCT acquisition protocols. Overall, these results demonstrate consistent agreement between 3D-DXA and QCT across sex, age, ethnicity, geographic regions, and clinical profiles. Taken together, these findings support the use of 3D-DXA as an accurate, non-invasive, and clinically accessible technology for advanced assessment of the cortical and trabecular compartments of the proximal femur.

## Introduction

Osteoporosis is a major public health concern worldwide due to its association with fragility fractures, particularly of the hip, which are linked to high morbidity, mortality, and healthcare costs [1], [2]. Thanks to advancements in osteoporosis medical management, the rate of fragility fracture among people aged 50 years and older has been decreasing in the last 20 years [3]. However, the social burden remains high, and the total number of fractures is projected to double by 2050 due to global ageing [4]. Dual-energy X-ray absorptiometry (DXA) is the standard clinical tool to assess bone mineral density (BMD) and derive T-scores for osteoporosis diagnosis [5]. Conventional DXA is a two-dimensional projection of the BMD, lacking the ability to differentiate between the two bone compartments (cortical and trabecular bone). T-scores derived from areal BMD (aBMD) set the standard thresholds to classify patients into osteopenic (T-score < -1) and osteoporotic (T-score < -2.5), although it was found that most fractures happen in osteopenic patients [6]. On the other hand, quantitative computed tomography (QCT) provides volumetric BMD and detailed three-dimensional (3D) structural parameters, but its widespread clinical use is limited by higher radiation dose, costs, and lack of standardization across scanners and protocols [7], [8].

3D reconstruction from DXA has been proposed by several groups to fill the gap between standard DXA and QCT, using a single-projection approach [9], [10], [11], or a multi-projection approach [12], [13]. The most common approach, developed independently by two groups, is based on statistical shape and density (or appearance) models (SSDMs) built from databases of QCT images. To reconstruct the patient-specific geometry and density distribution, the SSDM is optimized until its projection matches the DXA scan [9], [11].

The SSDM-based approach proposed by Väänänen et al., aims to reconstruct a proximal femur from a single DXA scan. It is built on two SSDMs of the proximal femur and pelvis. In an early study, the SSDMs were built from 34 cadaveric hip DXA scans (30 males and 4 females), and validation was performed on 12 in vivo hip DXAs from elderly women [11]. In subsequent studies, the SSDM was re-trained on 59 cadaveric femur QCT scans and validated against QCT in a separate group of 30 females [14], [15]. The calculation time for aligning the SSDM onto the DXA scan, however, was relatively high, reportedly 1.5 hour on a high-performance cluster [11].

The approach proposed by Humbert et al., known as *3D-DXA*, was designed for clinical use, achieving a computation time of under 2 minutes per scan, and positioned as a commercial solution (3D-Shaper® Software, 3D-Shaper Medical, Barcelona, Spain) [9]. 3D-DXA uses a SSDM of the proximal femur to derive cortical and trabecular bone parameters from a single DXA scan. The SSDM was built from a database of adults (>20 years old) from two clinical centers in Spain. The database included 111 healthy, osteopenic and osteoporotic subjects of both sexes. The accuracy of 3D-DXA was assessed against QCT in an independent dataset of 157 subjects from the same Spanish clinical centers.

Since the publication of its initial methodological and validation study [9], the software has been further developed to improve reconstruction performance, and regulatory clearances obtained in Europe, Japan and the United States (US), among other regions, have enabled its expanding clinical adoption. Thus, validation data across different populations and centers need to be reported to further assess the accuracy and the generalizability of the method, as also emphasized in a recent review [16].

Therefore, the aim of the present study is to report validation results comparing 3D-DXA and QCT parameters across four independent cohorts: the Spanish data used in previous work [9], two cohorts from Japan[17], and data from a multicenter US-based study [18].

## Methods

### Study subjects and medical images

This study included subjects from four cohorts who had both DXA and QCT acquisitions. The four cohorts are described as follows.

#### Adult population from Spain (same data used in earlier 3D-DXA validation study [9])

Study participants were recruited from two centers (Hospital de la Santa Creu i San Pau and CETIR ASCIRES, Barcelona, Spain). Exclusion criteria included skeletal diseases other than osteoporosis (e.g., osteoarthritis or Paget’s disease) and history of previous hip fracture. DXA images were acquired at CETIR ASCIRES using a Lunar iDXA scanner (GE Healthcare, Madison, WI) and at Hospital de la Santa Creu i Sant Pau using a Discovery W scanner (Hologic Inc., Waltham, MA). QCT-scans were performed at CETIR Esplugues PET (Esplugues de Llobregat, Spain) using a Philips Gemini GXL 16 (Philips Healthcare, Best, The Netherlands) and at CETIR Clínica Pilar Sant Jordi RM (Barcelona, Spain) using a Discovery CT750 HD scanner or a HiSpeed QX/I scanner (GE Healthcare) and a liquid K_2_HPO_4_ calibration phantom (Mindways Software Inc., Austin, TX).

#### Post-menopausal female population from the United States (Multi-Center Cohort)

Subjects from the Parathyroid Hormone and Alendronate (PaTH) study were included in the present analysis [18], [19]. The PaTH study included treatment-naive postmenopausal women who met at least one of the following criteria: T-score < −2.5; T-score < −2.0 combined with a previous fracture; or T-score < −2.0 and age > 65 years. The study was designed to evaluate the skeletal effects of parathyroid hormone therapy in osteoporosis. Participants were recruited from four clinical centers across the United States (Bangor, ME; Minneapolis, MN; New York, NY; and Pittsburgh, PA). DXA scans were acquired across all centers using Hologic Delphi or QDR 4500A and QCT scans were acquired in all centers with a GE CT-9800 using a solid hydroxyapatite (HA) phantom (Image Analysis, University of California, San Francisco, USA). For the present analysis, we included the patients with both DXA and QCT acquisitions available and only analyzed the baseline scans.

#### Osteoarthrosis population from Japan

The osteoarthrosis (OA) Japanese cohort was derived from a single-center study conducted at the Kawasaki Medical School between 2010 and 2012, as described in previous publication [17]. The patients were evaluated for OA and scheduled for hip replacement surgery. Hips affected by OA or previous surgery were excluded from the current analysis. DXA scans were acquired using a Hologic Discovery A densitometer and QCT scans were acquired with a GE Lightspeed Ultra 16 using a solid HA calibration phantom (B-MAS200, Kyoto Kagaku, Kyoto, Japan).

#### Young population from Japan

Healthy young volunteers recruited at Kawasaki Medical School in 2008 were included in the present study. DXA scans were acquired using a Hologic QDR Discovery A densitometer and QCT scans were acquired with a GE Lightspeed Ultra 16 using a solid HA calibration phantom (B-MAS200).

The study was approved by the Institutional Review Board or ethics committee at each clinical Center and written informed consent was obtained from all study subjects.

### DXA and 3D-DXA analysis

DXA scans from Hologic scanners were analyzed using APEX™ Software, and scans from GE healthcare were analyzed using enCORE software. Total hip and femoral neck areal BMD T-scores were calculated using NHANES III normative data, according international guidelines [5]. The lowest value between the total hip and femoral neck areal BMD T-score was used to classify the subjects in three categories: very low density (T-score < -2.5), low density (−2.5 < T-score < −1) and normal density (T-score > -1).

3D-DXA analysis was performed using 3D-Shaper® software version 2.14 (3D-Shaper Medical, Barcelona, Spain). 3D-Shaper software uses a SSDM generated from a database of QCT scans of the proximal femur [9]. An iterative optimization process is used to register the SSDM with the DXA scan. At each iteration, the projection of the SSDM is compared with the DXA image. The SSDM is optimized along its principal modes of variation to maximize the similarity between the projection and the DXA scan. This process, illustrated in Figure 1, yields a subject-specific, QCT-like 3D model of the proximal femur. Cortical and trabecular compartment are subsequently segmented using a model-based approach [20]. The clinical version of the 3D-Shaper software (v2.14) provides three output parameters calculated at the total femur: integral volumetric bone mineral density (vBMD, mg/cm^3^), trabecular vBMD (mg/cm^3^), and cortical surface bone mineral density (sBMD, mg/cm^2^).

**Figure 1.**
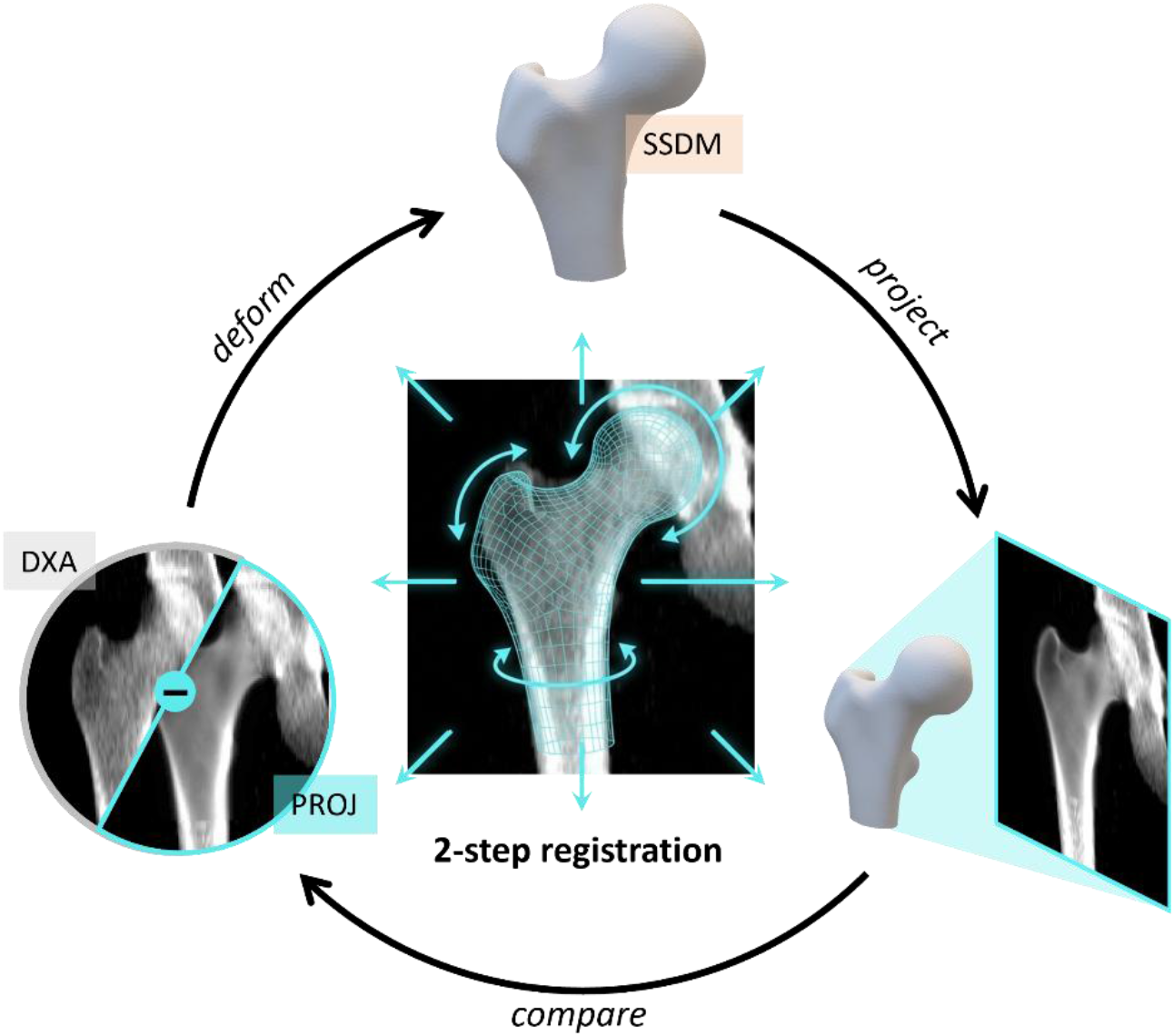
3D-DXA modeling iterative process. In a 2-step registration process, rigid and non-rigid transformations are applied to the statistical shape and density model (SSDM) to maximize the similarity between the dual-energy X-ray absorptiometry (DXA) scan and the projection (PROJ) of the SSDM. The final output is the subject-specific 3D SSDM.

### GE to Hologic conversion

To enable pooling of the Spanish dataset, whose data were collected using different dual-energy X-ray absorptiometry (DXA) manufacturers, conversion equations were used to transform 3D-DXA measurements derived from GE Healthcare hip DXA scans into Hologic-equivalent measurements. These conversion equations were previously derived from an independent dataset of 120 adult men and women (aged 22 to 75 years), as reported in a previous study [21]. Each subject underwent DXA imaging with both a Lunar iDXA (GE Healthcare, CETIR ASCIRES, Barcelona, Spain) and a Discovery scanner (Hologic, Hospital de la Santa Creu i Sant Pau, Barcelona, Spain). Linear regression analysis yielded the following conversion equations:

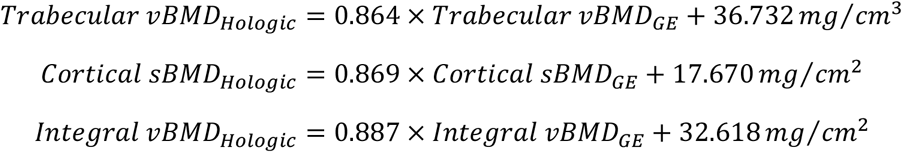

### QCT scans analysis

The QCT scans were processed according to a previously described methodology [9]. The CT images were calibrated using the phantoms scanned with the patients, and following recommendations of the manufacturers. To account for the differences between calibrations obtained using solid HA and liquid K_2_HPO_4_ phantoms, the following law was used to convert the density values (*vBMD*_*SOLID*_) of QCT volumes calibrated using solid phantoms (US and Japanese cohorts), into liquid phantom-equivalent density values (*vBMD*_*LIQUID*_) [22]:

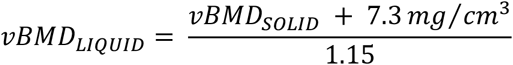

This conversion enabled consistent alignment between the QCT datasets and the 3D-DXA-derived measurements, as the software generates outputs in liquid phantom-equivalent units based on the calibration of its original training data.

The proximal femur ipsilateral to the DXA acquisition was segmented using a previously described semi-automatic thresholding approach [9]. Subsequently, the cortex was segmented using the same model-based approach as the one used by 3D-DXA [20]. To allow for comparison between 3D-DXA and QCT, the 3D-DXA geometries were rigidly aligned on the QCT geometries using the coherent point drift algorithm [23] and the regions of interest defined on the 3D-DXA data were projected onto the QCT geometries. Integral vBMD, trabecular vBMD, and cortical sBMD were calculated at the total femur, providing parameters equivalent to those obtained with 3D-DXA for comparison purposes.

#### Statistical Analysis

Accuracy results were reported as mean difference, as an estimate of the bias or systematic error, standard deviation (SD) of differences, an estimate of the random error, and Pearson’s correlation coefficient (R). Agreement between 3D-DXA and QCT was assessed using linear regression and Bland–Altman analysis, reporting mean difference and 95% limit of agreement (LoA), calculated as ±1.96 SD. Statistical significance was set at p < 0.05.

## Results

### Subjects Characteristics

The demographics of each cohort, as well as the DXA-derived BMD are reported in Table 1.

**Table 1.** Demographics and DXA-derived areal bone mineral density data for each cohort.

|  | Spain (N=97) | US (N=128) | Japan OA (N=205) | Japan Young (N=107) |
| --- | --- | --- | --- | --- |
| Age | 59.9 (13.4) | 69.9 (6.3) | 69.6 (10.5) | 21.8 (2.5) |
| Sex | 57F, 40M | 128F | 184F, 21M | 63F, 44M |
| Race/Ethnicity | 97W | 123W, 1A, 1B, 2O | 205A | 107A |
| BMI (kg/m<sup>2</sup>) | 25.7 (4.5) | 25.0 (4.1) | 25.5 (4.2) | 21.7 (2.6) |
| aBMD TH (g/cm<sup>2</sup>) | 0.88 (0.20) | 0.72 (0.08) | 0.76 (0.14) | 1.07 (0.16) |
| aBMD Neck (g/cm<sup>2</sup>) | 0.79 (0.17) | 0.61 (0.07) | 0.61 (0.12) | 0.93 (0.18) |

*Continuous variables are presented as mean (standard deviation)*
*F=female, M=Male; W=White, A=Asian, B=Black, O=Other; OA=Osteoarthritis; BMI=Body Mass Index, aBMD=areal Bone Mineral Density, TH=Total Hip.*

The Spanish cohort included 97 White subjects (40 male), aged 30–92 years. Based on T-score classification, 28 subjects had very low bone density, 34 had low bone density and 35 had normal bone density. For each subject, the DXA and QCT scans were acquired on the same day.

The US cohort from the PaTH study included 128 post-menopausal women aged 55–77 year, mainly White (1 Asian, 1 Black and 2 of other ethnicities). 40 subjects had very low bone density, 85 had low bone density, and 3 had normal bone density. DXA and QCT acquisitions were performed on the same day for 94% of the cohort. For the remaining participants, the time interval between scans did not exceed 11 days, with the exception of a single subject (62 days).

The Japanese OA cohort included 205 Asian adults (21 male) aged 40–90 years. 87 subjects had very low bone density, 89 had low bone density, and 29 had normal bone density. The mean time interval between the two acquisitions was 27 days (range: 2–390 days).

The Japanese Young cohort included 107 young Asian individuals (44 male) aged 19–33 years, all with normal bone density. All participants underwent both hip DXA and QCT imaging on the same day.

The details of the DXA and QCT scanners, the imaging parameters, and QCT calibration phantoms employed in this study are summarized in Table 2. All the DXA scans were acquired with Hologic DXA scanners except for a subset of the Spanish data (60 patients), acquired with a GE Lunar iDXA scanner. The QCT scans were acquired together with a phantom (either liquid or solid) in order to convert the voxel values (in Hounsfield Units, HU) to density values (in mg/cm^3^). Each cohort was acquired with a different CT scanner model, either from Philips or from GE Healthcare. The energy dose used for the acquisition was 120 kVp for all data except for the US data (80 kVp). Pixel size ranged between 0.68 mm × 0.68 mm (Japanese cohorts) and 1.1 mm × 1.1 mm (Spanish cohort), while the slice thickness ranged between 1 mm (Spanish cohort) and 3 mm (US cohort).

**Table 2.** Specifications of dual-energy X-rays absorptiometry (DXA) and quantitative computed tomography (CT) acquisitions for each cohort.

|  | DXA Scanners | CT Scanner Models | Energy/<br>Current | Pixel size<br>(mm x mm) | Slice thickness<br>(mm) | Calibration<br>Phantom |
| --- | --- | --- | --- | --- | --- | --- |
| Spanish data | Lunar iDXA<sup>1</sup> (38%);<br>Discovery W<sup>2</sup> (62%). | Philips Gemini GXL<sup>4</sup> (39%);<br>Discovery CT750 HD<sup>1</sup> (10%);<br>HiSpeed QX/I<sup>1</sup> (11%). | 120 kVp/<br>(138-188 mA) | 0.7 x 0.7 -<br>1.1 x 1.1 | 1.0 - 1.25 | Liquid:<br>Mindways<sup>5</sup> |
| US data | Delphi W<sup>2</sup> (19%);<br>QDR 4500A<sup>2</sup> (41%);<br>QDR 4500W<sup>2</sup> (40%). | CT-9800<sup>1</sup> | 80 kVp/<br>280 mA | 0.94 x 0.94 -<br>0.98 x 0.98 | 3.0 | Solid: Image<br>Analysis<sup>6</sup> |
| Japanese OA | Discovery A<sup>2</sup>. | Lightspeed Ultra 16<sup>1</sup> | 120 kVp/<br>250mA | 0.68 x 0.68 | 2.5 | Solid:<br>B-MAS200<sup>7</sup> |
| Japanese Young | Discovery A<sup>2</sup>. | Discovery STE<sup>1</sup> | 120 kVp/<br>250mA | 0.68 x 0.68 | 1.25 | Solid:<br>B-MAS200<sup>7</sup> |

<sup>1</sup>GE Healthcare, Madison, WI, USA; <sup>2</sup>Hologic Inc., Bedford, MA, USA; <sup>4</sup>Philips Healthcare, Best, The Netherlands; <sup>5</sup>Mindways Software Inc., Austin, TX, USA; <sup>6</sup>Image Analysis, University of California, San Francisco, CA, USA; <sup>7</sup>Kyoto Kagaku, Kyoto, Japan

#### Accuracy

Table 3 reports the mean and SD of 3D-DXA and QCT measurements, as well as mean and SD of the differences between 3D-DXA and QCT measurements, and correlation coefficients^1^. Mean difference, or bias, between the two methods was significant (p<0.001) for all parameters. The SD of the differences, which estimates the random error between the two methods, was 19.34 mg/cm^3^ for integral vBMD, 17.18 mg/cm^3^ for trabecular vBMD and 10.78 mg/cm^2^ for cortical sBMD. Strong correlations were found with coefficients ranging between 0.93 and 0.96.

**Table 3.** Accuracy results for all datasets combined.

|  | All (N=537) |  |  |  |
| --- | --- | --- | --- | --- |
|  | QCT:<br>Mean ± SD | 3D-DXA:<br>Mean ± SD | Difference<br>Mean ± SD | R |
| Integral vBMD<br>(mg/cm<sup>3</sup>) | 288.93 ± 71.28 | 281.35 ± 69.46 | -7.59*** ± 19.34 | 0.96 |
| Trabecular vBMD<br>(mg/cm<sup>3</sup>) | 157.95 ± 59.09 | 161.88 ± 52.31 | 3.93*** ± 17.18 | 0.96 |
| Cortical sBMD<br>(mg/cm<sup>2</sup>) | 139.12 ± 29.59 | 137.36 ± 29.77 | -1.76*** ± 10.78 | 0.93 |

<sup>1</sup>For consistency, 3D-DXA measurements from the Spanish cohort acquired using GE Healthcare DXA scanners were converted to Hologic-equivalent values using the conversion equations described in the Methods section, so that the combined data reported in Table 3 and Figure 2 include only Hologic or Hologic-equivalent measurements.
QCT = quantitative computed tomography; 3D-DXA = Three-dimensional dual-energy X-ray Absorptiometry; SD = standard deviation; v/s BMD = volumetric/surface bone mineral density; R = Pearson's correlation coefficient. Statistically significant differences reported as \*= $p < 0.05$ ; \*\*= $p < 0.01$ ; \*\*\*= $p < 0.001$ . All correlation coefficients were statistically significant ( $p < 0.001$ ).

Figure 2 presents the linear regression and Bland-Altman plots assessed for all data. Data from Spain, US, Japan OA and Japan Young are plotted in different colors to illustrate the performance of 3D-DXA vs QCT across different datasets. Bland-Altman plots are reported separately for each cohort in Supplementary Material (Figures Figure S1,Figure S2 andFigure S3).

**Figure 2.**
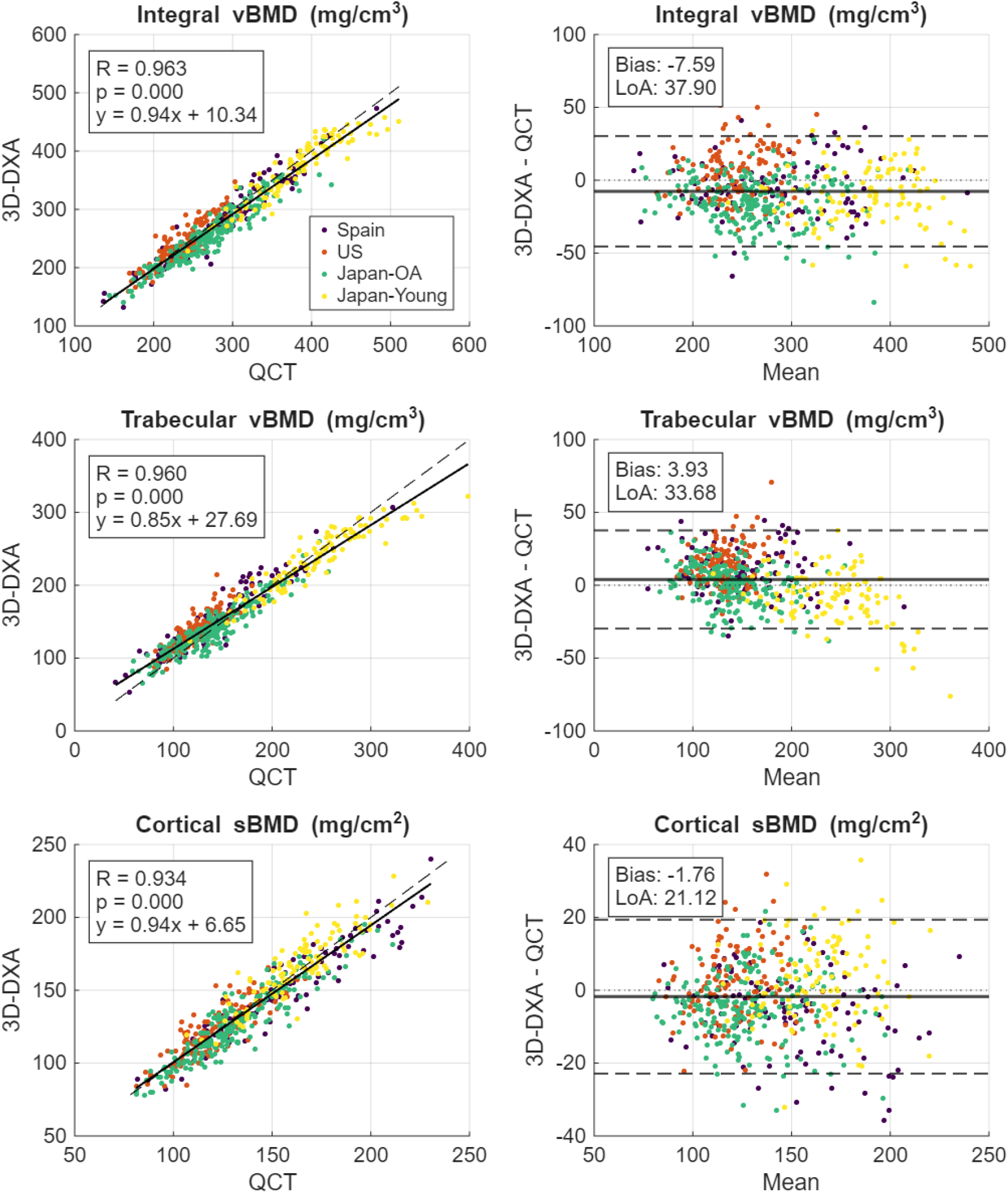
Linear regression (left) and Bland-Altman (right) plots for integral volumetric (v)BMD (top), trabecular vBMD (middle), and cortical surface (s)BMD (bottom). In linear regression plots, the solid line represents the regression line, while the dashed line represents the line of identity (y=x). In Bland-Altman plots, the x-axis represents the mean of the two measurements and the y-axis represents their difference. The solid lines represent mean difference (bias) and limits of agreements.

Table 4 presents the Spanish data stratified by DXA manufacturer. Accuracy is reported as mean difference, SD of differences and correlation coefficient. The correlations were strong in all groups, with correlation coefficients above 0.94. The GE Healthcare dataset showed significant (p<0.05) negative biases for trabecular vBMD and cortical sBMD, indicating an underestimation of bone density compared to QCT. In the Hologic cohort, all parameters presented a negative bias except trabecular vBMD. Overall, larger biases were observed for trabecular vBMD, whereas integral vBMD showed smaller biases. The conversion of GE 3D-DXA data to Hologic-equivalent measurements resulted in biases consistent with those observed in the Hologic dataset. This supports the use of Hologic-equivalent measurements when presenting pooled data, as in Table 3, Figure 2, and in the subsequent section of this article.

**Table 4.** Accuracy results within the Spanish dataset across the different manufacturers.

| Spain | GE (N=60) |  | GE (Hologic equivalent) (N=60) |  | Hologic (N=37) |  |
| --- | --- | --- | --- | --- | --- | --- |
| | Difference<br>Mean ± SD | R | Difference<br>Mean ± SD | R | Difference<br>Mean ± SD | R |
| Integral vBMD (mg/cm<sup>3</sup>) | -4.88 ± 24.31 | 0.96 | -4.16 ± 20.24 | 0.96 | -6.77* ± 15.67 | 0.97 |
| Trabecular vBMD (mg/cm<sup>3</sup>) | -6.06* ± 20.22 | 0.94 | 11.66*** ± 16.98 | 0.94 | 8.67*** ± 12.70 | 0.97 |
| Cortical sBMD (mg/cm<sup>2</sup>) | -3.91* ± 11.98 | 0.94 | -5.58*** ± 12.05 | 0.94 | -5.59** ± 10.10 | 0.96 |

QCT = Quantitative Computed Tomography; 3D-DXA = Three-dimensional dual energy x-ray absorptiometry; SD = Standard Deviations; v/sBMD = volumetric/surface Bone Mineral Density. Statistically significant biases are reported as \*= $p<0.05$ ; \*\*= $p<0.01$ ; \*\*\*= $p<0.001$ . All correlation coefficients were statistically significant with $p<0.001$ .

Details of the accuracy results for all cohorts included in the current study are presented in Table 5. Integral vBMD consistently showed the highest correlation coefficients, with values ranging from 0.87 to 0.96. Trabecular vBMD and cortical sBMD also demonstrated strong correlations, with correlation coefficients ranging from 0.82 to 0.95 and from 0.85 to 0.95, respectively. Bias for integral vBMD was significant in all cohorts and mostly negative, with values ranging between -14.84 mg/cm^3^ and 4.50 mg/cm^3^. For trabecular vBMD, bias was significant except for the Japanese OA cohort and ranged between -8.31 mg/cm^3^ and 14.41 mg/cm^3^. The bias for cortical sBMD was always significant except for the US cohort and ranged between -5.58 mg/cm^3^ and - 3.21 mg/cm^3^. The SD of differences, which estimates the random errors, was relatively stable across cohorts, ranging between 16.55 and 19.91 mg/cm^3^ for integral vBMD, between 13.52 and 18.47 mg/cm^3^ for trabecular vBMD and between 9.13 and 11.37 mg/cm^2^ for cortical sBMD.

**Table 5.** Accuracy results across all datasets (Spanish data including Hologic data and GE-converted).

|  | Spain (N=97) |  | US (N=128) |  | Japan OA (N=205) |  | Japan Young (N=107) |  |
| --- | --- | --- | --- | --- | --- | --- | --- | --- |
| | Difference<br>Mean ± SD | R | Difference<br>Mean ± SD | R | Difference<br>Mean ± SD | R | Difference<br>Mean ± SD | R |
| Integral vBMD (mg/cm<sup>3</sup>) | -5.16** ± 18.59 | 0.96 | 4.50** ± 17.42 | 0.87 | -14.84*** ± 16.55 | 0.95 | -10.35*** ± 19.91 | 0.92 |
| Trabecular vBMD (mg/cm<sup>3</sup>) | 10.52*** ± 15.49 | 0.95 | 14.41*** ± 13.52 | 0.82 | 0.66 ± 13.90 | 0.93 | -8.31*** ± 18.47 | 0.93 |
| Cortical sBMD (mg/cm<sup>2</sup>) | -5.58*** ± 11.29 | 0.95 | 1.53 ± 9.81 | 0.85 | -4.59*** ± 9.13 | 0.93 | 3.21** ± 11.37 | 0.87 |

OA = Osteoarthritis; SD = standard deviation; v/s BMD = volumetric/surface bone mineral density; R = Pearson's coefficient. Statistically significant biases reported as \*= $p<0.05$ ; \*\*= $p<0.01$ ; \*\*\*= $p<0.001$ . All correlation coefficients were statistically significant ( $p<0.001$ ).

To account for potential differences related to baseline bone density, the accuracy analysis was repeated including only subjects with very low bone density (T-score < −2.5), and the results are presented in Table 6. Since the Japanese Young cohort only included healthy subjects, the results from this cohort were excluded. Correlation coefficients in the very low density subsets were lower compared to the full datasets, due to the reduction of the range of density values. The highest decrease in correlation coefficients was observed for trabecular vBMD, assessed in the Spanish cohort, which was 0.95 using the full dataset, compared to 0.77 the very low density subset. All the other correlation coefficients were equal or larger than 0.8, indicating strong correlations. The US and Japanese cohorts consistently showed higher correlation coefficients for all parameters, compared to the Spanish cohort. Bias was consistent across different cohorts, with negative bias for integral vBMD bias (-13.37 mg/cm^3^ — -1.88 mg/cm^3^) and for cortical sBMD (-5.00 mg/cm^2^ — -1.49 mg/cm^2^), and with positive bias for trabecular vBMD (4.54 mg/cm^3^ — 11.38 mg/cm^3^). Random errors (SD of differences) remained within the same ranges as in the full cohorts for integral vBMD (14.05 mg/cm^3^— 19.18 mg/cm^3^) and trabecular vBMD (12.56 mg/cm^3^ — 17.59 mg/cm^3^) in all cohorts, while smaller random errors were found for cortical sBMD (7.43 mg/cm^2^— 8.76 mg/cm^2^).

**Table 6.** Accuracy results across the different dataset in subjects with very low bone density (T-score < −2.5).

|  | Spain (N=28) |  | US (N=40) |  | Japan (N=87) |  |
| --- | --- | --- | --- | --- | --- | --- |
|  | Difference<br>Mean ± SD | R | Difference<br>Mean ± SD | R | Difference<br>Mean ± SD | R |
| Integral vBMD<br>(mg/cm<sup>3</sup>) | -7.88* ± 19.18 | 0.85 | -1.88 ± 14.05 | 0.91 | -13.37*** ± 15.06 | 0.91 |
| Trabecular vBMD<br>(mg/cm<sup>3</sup>) | 9.17* ± 17.59 | 0.77 | 11.38*** ± 12.56 | 0.80 | 4.54** ± 14.77 | 0.80 |
| Cortical sBMD<br>(mg/cm<sup>2</sup>) | -2.53 ±<br>8.76 | 0.81 | -1.49 ± 7.59 | 0.90 | -5.00*** ± 7.43 | 0.87 |

SD = standard deviation; v/s BMD = volumetric/surface bone mineral density; R = Pearson's coefficient. Statistically significant biases reported as \*=*p*<0.05; \*\*=*p*<0.01; \*\*\*=*p*<0.001. All correlation coefficients were statistically significant (*p*<0.001).

## Discussion

This study assessed 3D-DXA against QCT across cohorts from different geographic regions, ethnicities, age groups, and clinical profiles. 3D-DXA has been proposed as a clinically applicable, low-radiation alternative to QCT to estimate volumetric compartment-specific BMD from standard DXA scans. This technology uses a SSDM generated from a database of QCT scans of the proximal femur from a White population from Spain. An independent database from the same population was used for validation as part of the original developments [9]. Given the widespread use of this technology, it was necessary to assess the accuracy of 3D-DXA beyond the original population.

3D-DXA-derived integral vBMD, trabecular vBMD and cortical sBMD bone parameters were found to strongly correlate with QCT across all cohorts assessed (R=0.82 – 0.96, Table 5).

Our findings align with and extend a previous validation study on a Spanish cohort of 157 subjects, reporting correlation coefficients of 0.96 (GE) and 0.95 (Hologic) for integral and 0.86 (GE) and 0.85 (Hologic) for trabecular vBMD [9]. Our study reanalyzed the same data with the latest version of the software (2.14), and found similar or greater correlations. In particular, for trabecular vBMD, the correlation coefficients increased to 0.94 (GE) and 0.97 (Hologic) (Table 5). At the same time, the random errors (SD of differences) in these cohorts decreased slightly. In addition, the current findings support the use of 3D-DXA technology with data from different race/ethnic groups (White and Asian), and different countries (US, and Japan).

Relying solely on correlation coefficient to assess accuracy of density measurements has notable limitations, as the value naturally decreases if the range of density values is reduced. This dependency was evident when evaluating the full cohorts: the lowest overall correlation coefficients were found in the US-based dataset which was exclusively composed of postmenopausal women with low bone mass or osteoporosis (Table 5). However, when the comparison was restricted to only include very low bone density subjects, correlation coefficients were consistent across all cohorts, with the US-based dataset even showing slightly higher correlation coefficients comparted to the Spanish dataset (Table 6). Thus, the following sections will discuss the biases and random errors as additional metrics to evaluate 3D-DXA accuracy.

A previous ex-vivo study evaluated 3D-DXA against QCT scans using cadaveric femurs scanned with a GE Healthcare DXA scanner [24]. A strong correlation (R^2^=0.93) and a random error of 21 mg/cm^3^ were reported for integral vBMD. These values align with those reported for the GE data included in the current study (R=0.96, resulting in R^2^=0.92, and a random error of 24.35 mg/cm^3^, Table 4). However, this ex-vivo study reported a higher bias (−64 mg/cm^3^) than in our study (- 4.88 mg/cm^3^, non-significant, Table 4). As noted by the authors, this discrepancy may be partly explained by the absence of a correction applied to their QCT data, acquired using a solid phantom, to convert them into liquid-equivalent values, as was done in the present study. This approach introduces a methodological bias by comparing solid-phantom-calibrated QCT measurements directly against 3D-Shaper outputs, which are inherently expressed in liquid-phantom-equivalent units. Another factor that may have affected the accuracy results reported by the authors is the use of different CT segmentation methods than those employed by the 3D-Shaper software and used to process the CT data in the present study.

In our study, the presence of significant biases indicates a systematic, although moderate, difference in the QCT and 3D-DXA measurements. In particular, when pooling the data from all cohorts together, trabecular vBMD was systematically overestimated (up to 3.93 mg/cm^3^) while integral vBMD and cortical sBMD were underestimated (-7.59 mg/cm^3^ and -1.76 mg/cm^2^ respectively, as reported in Table 3).

Our results further indicate that the bias varies across the different cohorts. This could potentially arise from the different DXA scanners used in the studies. Most of the DXA images were acquired from Hologic scanners, with the exception of a subset from the Spanish cohort. To ensure comparability, parameters derived from the GE scanner were converted using previously established equations. This harmonization resulted in biases that were more consistent with those observed in the Hologic dataset (Table 4). Previous works showed that using different scanner models from the same manufacturer yields comparable results[25], [26]. In GE scanners, this inter-scanner variability study reported biases below 3.0 mg/cm^3^ for integral vBMD, 0.9 mg/cm^2^ for cortical sBMD and 2.7 mg/cm^3^ for trabecular vBMD [25]. For Hologic scanners, no statistically significant bias was observed across different models, with biases below 2.5 mg/cm^3^ for integral vBMD, 1.7 mg/cm^2^ for cortical sBMD and 2.9 mg/cm^3^ for trabecular vBMD [26]. Since inter-scanner variability is substantially lower than the variation in bias observed across the validation cohorts in the present study, we hypothesize that scanner model or manufacturer has only a limited impact and likely explains these variations only marginally.

The cohort profile could also potentially influence the observed bias. The cohorts included in this study included different sexes, ethnicities, and individual profiles from young subjects to patients with osteoporosis or osteoarthritis. Interestingly, the bias in the Japanese OA and Japanese Young were rather similar, as shown in Table 5 and Figure 1, despite marked differences in subject characteristics between these groups, as reported in Table 1. This suggests that cohort profile may contribute only minimally to the differences in bias observed across the cohorts in this study.

Finally, differences in QCT scanners, acquisition protocols, and calibration phantoms (solid versus liquid) may also have influenced the observed bias. Six scanners were used, with substantial variation in acquisition parameters, including in-plane pixel size (0.68 mm × 0.68 mm to 1.1 mm × 1.1 mm) and slice thickness (1–3 mm). For example, the higher bias observed for trabecular vBMD in the US cohort may be partly attributable to lower image resolution, characterized by larger pixel size and slice thickness compared with the other cohorts. Notably, the Japanese OA and Japanese Young cohorts had the most similar QCT acquisition characteristics and used the same calibration phantom, and they also exhibited similar biases. This supports the hypothesis that differences in QCT scanners, protocols, and phantoms are likely a major factor underlying the variability in bias observed across the evaluated cohorts. Further validation studies are needed to better characterize the biases associated with differences in QCT acquisition protocols.

While assessing systematic bias is a critical component of any accuracy study, bias can generally be corrected to align the measurements with a validated ground truth. Random error, conversely, represents the inherent and uncorrectable scatter of the measurements. Notably, the random errors, assessed as the SD of the differences or the LoA, remained consistent across cohorts, with SD of differences ranging between 16.55 and 19.91 mg/cm^3^ for integral vBMD, 13.52 and 18.47 mg/cm^3^ for trabecular vBMD, and 9.13 and 11.37 mg/cm^2^ for cortical sBMD. Furthermore, to account for potential differences related to baseline bone density, we evaluated subsets of subjects with very low bone density (T-score ≤ -2.5). In this very low bone density subset, random errors were comparable to those observed in the full cohorts, with SDs of the between 14.05 and 19.18 mg/cm^3^ for integral vBMD, 12.56 and 17.59 mg/cm^3^ for trabecular vBMD, and 7.43 and 8.76 mg/cm^2^ for cortical sBMD.

The present study also addresses several key concerns raised in a recent critical review of 3D-DXA [16], particularly those related to the accuracy of 3D-DXA-derived cortical measurements, the limited validation populations, the overreliance on correlation metrics, and the potential lack of generalizability. By including a large, multi-center cohort (N=537) spanning different ethnicities (White and Asian), geographic regions (Spain, the US, and Japan), age groups, and clinical profiles, this work helps mitigate concerns regarding population bias and the restricted training dataset of the 3D-DXA SSDM. In addition to reporting strong correlations with QCT (R=0.82–0.96), the study incorporates Bland–Altman analyses and detailed assessments of both systematic and random errors, thereby providing a more comprehensive evaluation of measurement accuracy, including for cortical parameters. Furthermore, the consistency of random errors across cohorts supports the robustness of the method across diverse populations.

One limitation of our study lies is the underrepresentation of certain racial and ethnic groups in the validation datasets, notably Black populations, with only two individuals included. Although 3D-DXA has previously been applied successfully in this population [27], further dedicated validation studies could be performed to support its applicability. Future research should explore the accuracy of 3D-DXA across various etiologies of secondary osteoporosis, including chronic kidney disease (CKD) and glucocorticoid-induced bone loss. Moreover, as our primary objective was to confirm clinical generalizability across new populations, our evaluation was limited to the parameters available in the clinical version of the 3D-Shaper software. A more extensive evaluation of research software metrics, such as cortical thickness and cortical vBMD, was outside the scope of this study, although these have been investigated elsewhere[9], [28].

## Conclusion

Overall, 3D-DXA–derived bone measurements showed strong agreement with those obtained from QCT. Random errors remained consistent across sex, age, geographic region, ethnicity, and clinical profile. The cohort-dependent biases observed are likely attributable to differences in acquisition protocols. Taken together, these findings support the use of 3D-DXA as an accurate, non-invasive, and clinically accessible technology for advanced assessment of cortical and trabecular compartments of the proximal femur.

## Data Availability

All validation results produced in the present study are available upon reasonable request to the authors

## Supplementary Material

**Figure S1.**
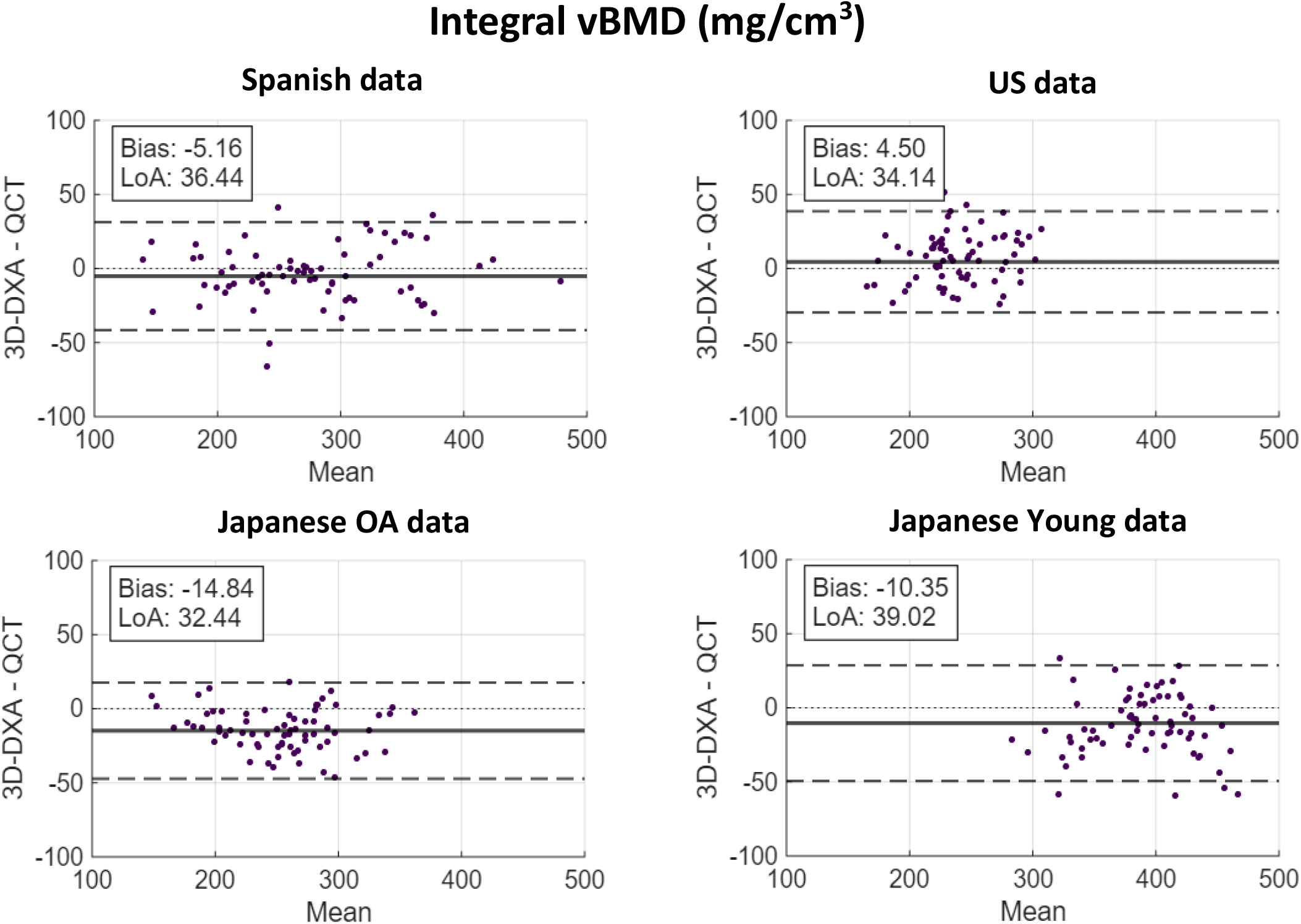
Bland-Altman plots for integral volumetric bone mineral density (vBMD) for the four cohorts in this study. The x-axis represents the mean of the two measurements and the y-axis represents their difference. The solid lines represent mean difference (bias) and limits of agreementsd (LoA).

**Figure S2.**
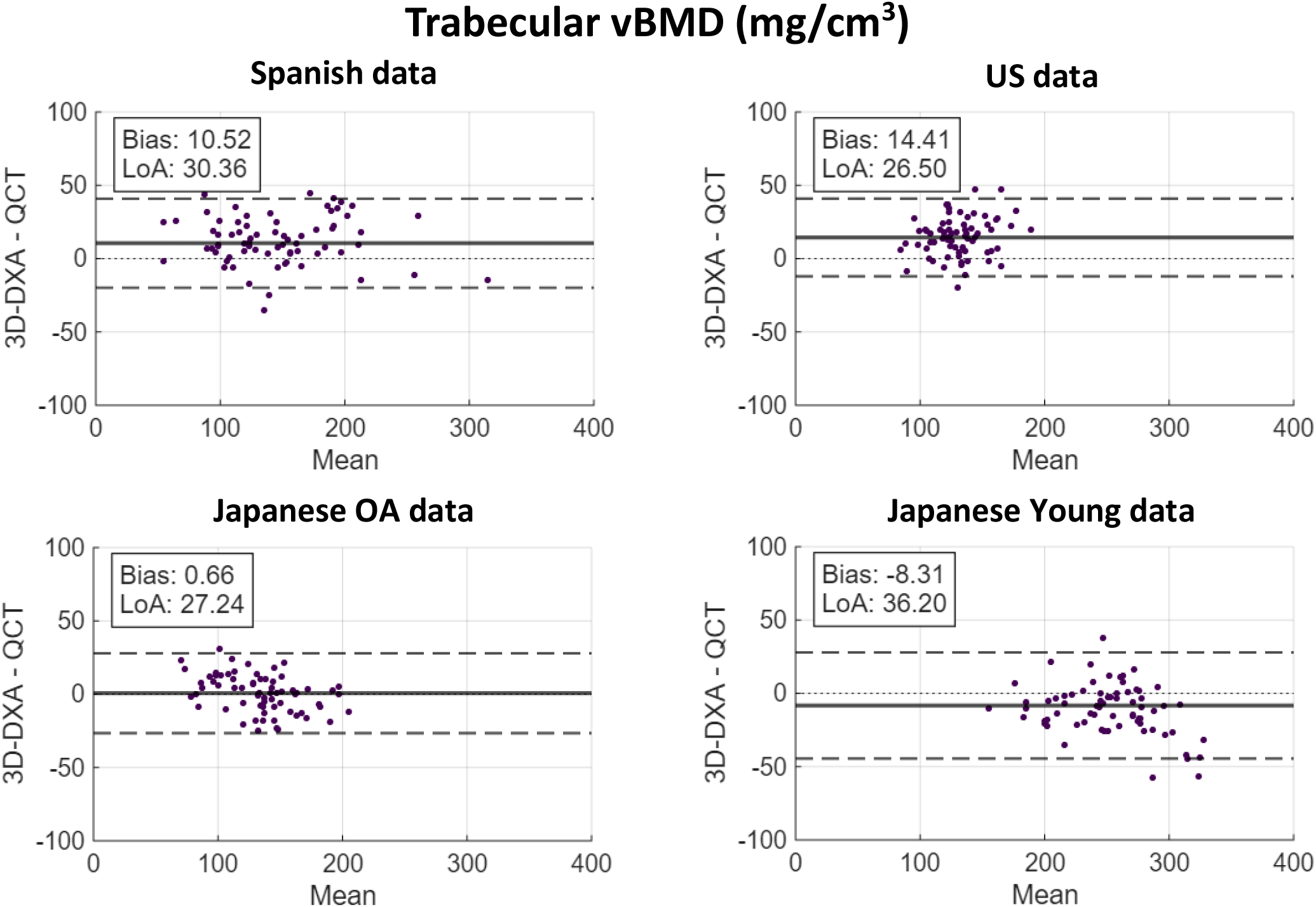
Bland-Altman plots for trabecular volumetric bone mineral density (vBMD) for the four cohorts in this study. The x-axis represents the mean of the two measurements and the y-axis represents their difference. The solid lines represent mean difference (bias) and limits of agreements (LoA).

**Figure S3.**
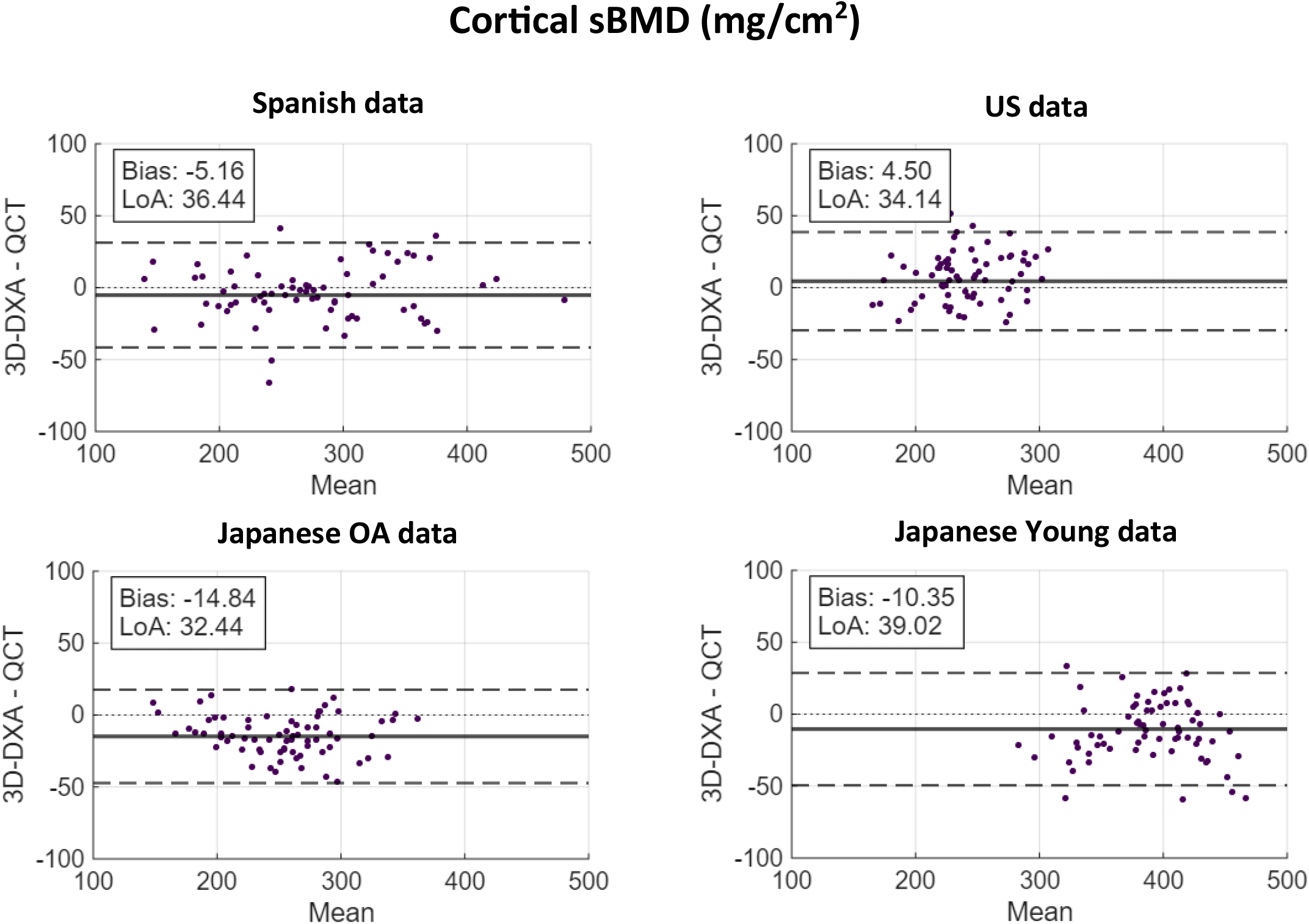
Bland-Altman plots for cortical surface bone mineral density (sBMD) for the four cohorts in this study. The x-axis represents the mean of the two measurements and the y-axis represents their difference. The solid lines represent mean difference (bias) and limits of agreements (LoA).

## Footnotes

1 For consistency, 3D-DXA measurements from the Spanish cohort acquired using GE Healthcare DXA scanners were converted to Hologic-equivalent values using the conversion equations described in the Methods section, so that the combined data reported in Table 3 and Figure 2 include only Hologic or Hologic-equivalent measurements.

## Notes

### Competing Interest Statement

Ludovic Humbert is an employee and shareholder of 3D-Shaper Medical. Marta I. Bracco is an employee of 3D-Shaper Medical. All the other authors have declared no competing interest.

### Funding Statement

This study did not receive any funding

### Author Declarations

The study protocol was reviewed and approved by the Institutional Review Boards (IRB) or Clinical Research Ethics Committees (CEIm) of all participating hospitals: Spanish centers: Hospital de la Santa Creu i Sant Pau and CETIR ASCIRES, Barcelona, Spain; USA centers: Bangor, ME; Minneapolis, MN; New York, NY; and Pittsburgh, PA; Japan center: Kawasaki Medical School. The study was conducted in accordance with the ethical standards of the Declaration of Helsinki. All participants provided written informed consent prior to inclusion in the study.

